# Modelling the impact of health-related variables, age, migration, and socio-economic factors in the geographical distribution of early tested case-fatality risks associated with COVID-19 in Mexico

**DOI:** 10.1101/2020.11.26.20239376

**Authors:** Ricardo Ramírez-Aldana, Juan Carlos Gomez-Verjan, Omar Yaxmehen Bello-Chavolla, Carmen García-Peña

## Abstract

COVID-19 is a respiratory disease caused by SARS-CoV-2, which has significantly impacted economic and public healthcare systems world-wide. SARS-CoV-2 is highly lethal in older adults (>65 years old) and in cases with underlying medical conditions including chronic respiratory diseases, immunosuppression, and cardio-metabolic diseases including severe obesity, diabetes, and hypertension. The course of the COVID-19 pandemic in Mexico has led to many fatal cases in younger patients attributable to cardio-metabolic conditions. Here, we aimed to perform an early spatial epidemiological analysis for the COVID-19 outbreak in Mexico to evaluate how tested case-fatality risks (t-CFRs) are geographically distributed and to explore spatial predictors of early t-CFRs considering the variation of their impact on COVID-19 fatality across different states in Mexico, controlling for the severity of the disease. As results, considering health related variables; diabetes and obesity were highly associated with COVID-19 fatality. We identified that both external and internal migration had an important impact over early COVID-19 risks in Mexico, with external migration having the second highest impact when analyzing Mexico as a whole. Physicians-to-population ratio, as a representation of urbanity, population density, and overcrowding households, has the highest impact on t-CFRs, whereas the age group of 10 to 39 years was associated with lower risks. Geographically, the states of Quintana Roo, Baja California, Chihuahua, and Tabasco had higher t-CFRs and relative risks comparing with a national standard, suggesting that risks in these states were above of what was nationally expected; additionally, the strength of the association between some spatial predictors and the COVID-19 fatality risks variates by zone depending on the predictor.

## INTRODUCTION

COVID-19 is a respiratory disease caused by SARS-CoV-2 (severe acute respiratory syndrome coronavirus 2), which has caused almost twenty million cases around the world and caused 790,000 deaths as of August 20^th^, 2020 (1). SARS-CoV-2 is a highly contagious RNA virus from the Coronaviridae family, with a small genome of about 30,000 nucleotides closely related to the bat coronavirus (RaTG13) (2). Given its global spread, there is an urgent need for scientific and health systems around the world to understand the epidemiology, pathogenicity, and mechanisms of immunological defenses to develop possible therapeutic and public health alternatives to fight against one of the most outstanding threats to public health since *Spanish Influenza* over a hundred years ago (3). According to the World Health Organization (WHO), the groups most susceptible to acquire infection and develop adverse outcomes are people with underlying medical conditions and older adults (>65 years old), particularly those living at nursing homes. Medical conditions which have been associated with increased susceptibility for adverse outcomes related to SARS-CoV-2 infection include chronic obstructive pulmonary disease (COPD), chronic kidney disease (CKD), cardiovascular diseases, liver diseases, moderate asthma, immunosuppression (HIV/AIDS, bone marrow transplantation, cancer treatment, and genetic immune deficiencies), and particularly, severe obesity, diabetes, and hypertension (4). These associations may be related to the strong link between pro-inflammatory cytokines in response to infection and the pathogenesis of SARS-CoV-2, which can be seen in pneumonia patients with severe COVID-19 disease exhibiting systemic hyper-inflammation known as cytokine storm or as a secondary hemophagocytic lymphohistocytosis (5).

On the other hand, accordingly to the OCDE, in the last ten years the countries with the highest prevalence of cardio-metabolic conditions linked to increased risk of severe COVID-19, including obesity, type 2 diabetes, and hypertension, are Mexico and the United States of America (USA), when considering adults from 15 to 74 years old (6). Thus, considering that the population pyramid is flattening more and more, indicating an aging population structure; the high rates of diabetes and obesity in Mexico are likely to increase susceptibility to higher rates of mortality attributable to COVID-19 even in younger populations (7).

In response to SARS-CoV-2 spread worldwide, mobility restrictions have been imposed to reduce community-level transmission; nevertheless, early influence of human mobility particularly internal and external migration has largely driven the spread of COVID-19 (8). When considering the high transmissibility of SARS-CoV-2, human mobility gains relevance in early stages of spread, in which people travelling from other countries can drive increased rates of transmission (9). SARS-CoV-2 spread related to human mobility is relevant, particularly considering that the risk of transmission and adverse outcomes are related to inequalities and suboptimal socio-economic conditions(10). In this sense, the risk of becoming infected and dying might increase in areas without optimum socio-economic conditions, for instance, spatial units in which people live in overcrowding households or without access to potable water or drainage.

Spatial analyses allow us to understand how the fatality risks are distributed along a territory, the presence of spatial clusters, and how the effects of the variables associated with the risks variate along any given territory. In this sense, several examples of geoepidemiology studies in autoinflammatory diseases, specific syndromes, infectious diseases, among others, have been helpful for the development of public health policies and to understand the structure of disease spread (11–13). In the present work, we performed spatial analyses in order to derive spatial relationships corresponding to the geographical phenomenon of the COVID-19 outbreak in Mexico and its associated fatality risks, using for this task statistical methods including spatial clustering through local indicators of spatial autocorrelation and generalized geographically weighted regression. Additionally, we wanted to identify spatial units or regions in Mexico that should be considered and analyzed with care to better understand the propagation of the disease and its associated fatality risks in the early stages of COVID-19 spread.

## MATERIAL AND METHODS

### Data sources

We obtained state-level variables considering the 32 states in Mexico (**Table 1**). A first group of variables were obtained from the epidemiological surveillance entity of Mexico (Direccion General de Vigilancia Epidemiologica, Secretaria de Salud) at an individual level, the data set corresponded to observations until April 21^st^, 2020 (14). These variables concern health and very general socioeconomic information associated with people who were suspected for COVID-19 in Mexico and underwent real time RT-PCR for SARS-CoV-2 confirmation. Available variables include the presence of diabetes, obesity, chronic renal problems (CKD), chronic obstructive pulmonary disease (COPD), pregnancy, hypertension, immunosuppression, cardiovascular disease, pneumonia, as well as age, and whether the patient was hospitalized, admitted to an intensive care unit (ICU), or required intubation. We grouped age into groups as explained in the model selection process below and we finally used three age groups: 10-39, 40-69, and 70 years old and over. We also computed the risk of death due to the disease on individuals who were tested for SARS-CoV-2, or tested case-fatality risks (t-CFRs), by considering as a death that which was recorded after having a positive test (the information concerning positive tests and death is available in the epidemiological data set), method consistent with the official numbers. We aggregated all variables at a state level as counts and used them as relative frequencies (rates) in all analyses.

**Table 1.** Features extracted for all the analyses by state used to predict tested case-fatality risks of COVID-19 in Mexico. Abbreviations: GDP, Gross Domestic Product; COPD, Chronic Obstructive Pulmonary Disease; and ICU, Intensive Care Unit.

| State | Deaths caused by<br>COVID-19* | Number<br>of tested<br>individuals<br>(offset)* | Rural 2006<br>(%)** | Poverty<br>2018 (%)<br>*** | Density<br>2015+ | Literacy<br>2015<br>(%)+ | GDP<br>2018<br>(%)+ | Temperature<br>March 2020<br>(°C)++ | Internal<br>migration<br>2014<br>(%)+ | External<br>migration<br>2014<br>(%)+ | Physicians<br>by 1000<br>people<br>2015+++ | Overcrowding<br>2017 (%)+ |
| --- | --- | --- | --- | --- | --- | --- | --- | --- | --- | --- | --- | --- |
| AGUASCALIENTES | 2 | 1070 | 18.84 | 26.18 | 233.70 | 97.00 | 1.31 | 19.30 | 3.60 | 0.70 | 1.33 | 3.90 |
| BAJA CALIFORNIA | 100 | 2353 | 6.99 | 23.26 | 46.40 | 97.60 | 3.14 | 14.40 | 3.80 | 1.90 | 1.05 | 6.40 |
| BAJA CALIFORNIA<br>SUR | 9 | 978 | 15.22 | 18.07 | 9.60 | 96.80 | 0.98 | 19.80 | 8.20 | 0.50 | 1.50 | 10.40 |
| CAMPECHE | 4 | 229 | 26.00 | 46.25 | 15.60 | 92.90 | 2.98 | 28.00 | 4.10 | 0.20 | 1.16 | 16.50 |
| COAHUILA DE<br>ZARAGOZA | 23 | 2472 | 9.95 | 22.49 | 19.50 | 97.10 | 3.50 | 22.10 | 2.10 | 0.40 | 1.44 | 4.50 |
| COLIMA | 2 | 133 | 12.41 | 30.87 | 126.40 | 95.60 | 0.61 | 26.60 | 5.60 | 1.50 | 1.56 | 10.70 |
| CHIAPAS | 4 | 440 | 52.26 | 76.41 | 71.20 | 84.20 | 1.56 | 25.60 | 1.50 | 0.50 | 0.53 | 20.50 |
| CHIHUAHUA | 36 | 705 | 15.51 | 26.28 | 14.40 | 95.00 | 3.20 | 16.70 | 1.20 | 1.20 | 1.11 | 5.60 |
| CIUDAD DE<br>MÉXICO | 224 | 10183 | 0.35 | 30.55 | 5967.30 | 97.70 | 17.64 | 19.40 | 2.10 | 0.40 | 2.99 | 7.10 |
| DURANGO | 4 | 407 | 32.77 | 37.35 | 14.20 | 96.20 | 1.13 | 18.40 | 2.60 | 0.80 | 1.17 | 5.40 |
| GUANAJUATO | 9 | 2482 | 30.29 | 43.38 | 191.30 | 93.00 | 4.11 | 20.60 | 1.50 | 1.00 | 0.84 | 5.50 |
| GUERRERO | 16 | 583 | 42.44 | 66.47 | 55.60 | 85.50 | 1.37 | 26.20 | 1.90 | 1.10 | 0.64 | 27.60 |
| HIDALGO | 16 | 608 | 47.69 | 43.85 | 137.30 | 91.00 | 1.54 | 20.50 | 3.90 | 0.90 | 0.66 | 8.20 |
| JALISCO | 15 | 3440 | 13.85 | 28.43 | 99.80 | 95.80 | 6.91 | 22.40 | 2.20 | 1.10 | 1.26 | 5.30 |
| MÉXICO | 71 | 5759 | 12.74 | 42.72 | 724.20 | 95.80 | 8.93 | 16.30 | 2.80 | 0.40 | 0.81 | 8.60 |
| MICHOACÁN | 19 | 1124 | 32.06 | 46.04 | 78.20 | 90.80 | 2.43 | 21.70 | 1.70 | 1.20 | 0.72 | 10.10 |
| MORELOS | 14 | 525 | 13.91 | 50.82 | 390.20 | 94.30 | 1.13 | 24.70 | 3.30 | 0.90 | 1.04 | 10.80 |
| NAYARIT | 5 | 299 | 33.48 | 34.77 | 42.40 | 94.30 | 0.68 | 24.70 | 4.30 | 1.40 | 1.06 | 8.10 |
| NUEVO LEÓN | 6 | 4482 | 5.64 | 14.53 | 79.80 | 97.40 | 7.47 | 22.90 | 2.70 | 0.40 | 1.30 | 3.10 |
| OAXACA | 7 | 522 | 52.89 | 66.35 | 42.30 | 84.20 | 1.48 | 24.60 | 2.00 | 1.20 | 0.61 | 16.90 |
| PUEBLA | 53 | 1651 | 29.38 | 58.92 | 179.80 | 90.80 | 3.39 | 20.00 | 2.30 | 0.60 | 0.83 | 9.70 |
| QUERÉTARO | 6 | 630 | 30.13 | 27.58 | 174.40 | 94.70 | 2.33 | 22.00 | 5.60 | 0.90 | 0.85 | 6.90 |
| QUINTANA ROO | 47 | 982 | 14.42 | 27.57 | 33.60 | 95.50 | 1.63 | 26.80 | 8.00 | 0.70 | 0.96 | 17.50 |
| SAN LUIS POTOSÍ | 5 | 1167 | 37.31 | 43.40 | 44.50 | 92.90 | 2.11 | 24.60 | 2.70 | 0.80 | 0.85 | 6.20 |
| SINALOA | 52 | 1580 | 29.17 | 30.88 | 51.70 | 95.20 | 2.23 | 22.90 | 2.50 | 0.70 | 1.16 | 9.20 |
| SONORA | 13 | 869 | 14.23 | 28.19 | 15.90 | 96.70 | 3.26 | 17.70 | 2.90 | 1.10 | 1.39 | 7.00 |
| TABASCO | 55 | 1266 | 44.98 | 53.65 | 96.90 | 93.80 | 2.61 | 27.60 | 2.30 | 0.30 | 0.92 | 12.00 |
| TAMAULIPAS | 7 | 1210 | 12.74 | 35.07 | 42.90 | 96.00 | 2.82 | 25.20 | 1.80 | 0.30 | 1.20 | 6.90 |
| TLAXCALA | 5 | 587 | 21.77 | 48.38 | 318.40 | 95.20 | 0.56 | 16.30 | 3.40 | 0.60 | 0.81 | 9.50 |
| VERACRUZ | 13 | 1520 | 39.33 | 61.78 | 113.00 | 89.80 | 4.59 | 23.70 | 3.00 | 0.60 | 0.87 | 12.60 |
| YUCATÁN | 12 | 959 | 17.01 | 40.80 | 53.10 | 91.90 | 1.46 | 26.90 | 3.60 | 0.40 | 1.39 | 13.90 |
| ZACATECAS | 3 | 419 | 42.73 | 46.76 | 21.00 | 94.90 | 0.88 | 19.10 | 2.10 | 1.40 | 0.89 | 5.00 |

\* Dirección General de Epidemiología, Secretaría de Salud (General Direction of Epidemiology, Health Ministry)
\*\* Instituto Nacional de Salud Pública, Encuesta Nacional de la Dinámica Demográfica (National Public Health Institute, National Survey of Demographic Dynamic)
\*\*\* Estimations obtained by CONEVAL (Consejo Nacional de Evaluación de la Política de Desarrollo Social, National Council for the Evaluation of Social Development Policy) based on the ENIGH (National Survey of Household Income and Expenditure) 2008, 2010, 2012, 2014, 2016, and 2018.

**Table 1(continued)**
| State | Diabetes (%)* | Obesity (%)* | Chronic renal problems (%)* | COPD (%)* | Pregnancy (%)* | Hypertension (%)* | Immuno-deficiency (%)* | Cardio-vascular (%)* | Pneumonia (%)* | ICU (%)* | Intubated (%)* | Hospitalized (%)* | Age group 10-39(%)* | Age group 40-69(%)* | Age group 70 years and over(%)* |
| --- | --- | --- | --- | --- | --- | --- | --- | --- | --- | --- | --- | --- | --- | --- | --- |
| AGUASCALIENTES | 7.216 | 14.204 | 1.947 | 1.833 | 0.573 | 13.402 | 1.833 | 2.864 | 7.446 | 0.458 | 0.916 | 12.257 | 54.296 | 33.333 | 4.582 |
| BAJA CALIFORNIA | 12.420 | 18.059 | 1.570 | 1.713 | 1.071 | 22.484 | 2.355 | 3.854 | 21.485 | 1.213 | 2.784 | 27.195 | 49.179 | 43.683 | 3.854 |
| BAJA CALIFORNIA SUR | 11.350 | 10.763 | 0.978 | 0.978 | 2.544 | 14.677 | 2.153 | 3.131 | 13.503 | 1.566 | 1.370 | 17.613 | 53.816 | 35.225 | 4.892 |
| CAMPECHE | 11.111 | 16.239 | 0.000 | 0.855 | 0.000 | 12.821 | 2.137 | 2.137 | 16.667 | 2.137 | 1.282 | 26.923 | 46.581 | 45.299 | 5.556 |
| COAHUILA DE ZARAGOZA | 11.965 | 16.079 | 2.015 | 1.847 | 0.756 | 16.037 | 2.603 | 2.981 | 8.942 | 1.847 | 0.672 | 14.484 | 47.397 | 44.249 | 5.584 |
| COLIMA | 14.388 | 19.424 | 5.036 | 2.878 | 2.158 | 15.827 | 2.158 | 3.597 | 25.899 | 2.878 | 2.878 | 33.094 | 52.518 | 34.532 | 7.194 |
| CHIAPAS | 11.836 | 13.050 | 1.973 | 2.883 | 1.669 | 15.630 | 2.276 | 2.731 | 17.299 | 3.642 | 3.035 | 27.314 | 49.469 | 39.150 | 4.856 |
| CHIHUAHUA | 15.385 | 13.501 | 2.826 | 4.710 | 1.413 | 21.821 | 3.297 | 5.181 | 33.438 | 9.733 | 3.925 | 45.997 | 45.997 | 40.816 | 7.849 |
| CIUDAD DE MÉXICO | 10.796 | 15.511 | 1.658 | 1.872 | 0.618 | 14.970 | 2.688 | 3.049 | 14.670 | 2.044 | 2.113 | 20.356 | 44.619 | 47.926 | 4.767 |
| DURANGO | 15.615 | 13.621 | 2.990 | 4.983 | 0.831 | 24.917 | 4.485 | 5.980 | 19.767 | 2.326 | 2.159 | 33.389 | 43.355 | 39.203 | 13.787 |
| GUANAJUATO | 10.131 | 11.997 | 1.908 | 3.942 | 1.399 | 14.455 | 2.755 | 2.671 | 10.428 | 1.696 | 1.526 | 17.338 | 50.996 | 34.718 | 6.867 |
| GUERRERO | 15.456 | 17.531 | 2.801 | 3.216 | 1.245 | 17.635 | 1.763 | 3.734 | 26.349 | 5.394 | 4.253 | 33.091 | 43.154 | 44.813 | 7.573 |
| HIDALGO | 18.491 | 15.976 | 4.142 | 4.734 | 1.479 | 19.970 | 4.438 | 2.071 | 21.893 | 1.627 | 2.959 | 31.953 | 42.751 | 44.231 | 9.172 |
| JALISCO | 10.876 | 13.425 | 3.533 | 3.625 | 1.475 | 16.559 | 2.888 | 3.840 | 12.442 | 1.874 | 1.260 | 20.399 | 49.124 | 37.665 | 7.957 |
| MÉXICO | 12.899 | 15.374 | 2.358 | 2.358 | 0.838 | 15.140 | 2.747 | 3.079 | 24.318 | 2.864 | 3.254 | 33.164 | 45.733 | 44.564 | 5.943 |
| MICHOACÁN | 13.541 | 16.003 | 2.752 | 4.272 | 1.376 | 19.406 | 2.462 | 4.345 | 22.230 | 2.172 | 2.462 | 29.616 | 41.999 | 41.202 | 11.369 |
| MORELOS | 14.397 | 14.786 | 2.918 | 4.086 | 1.167 | 17.899 | 2.724 | 2.529 | 24.708 | 4.086 | 3.502 | 34.825 | 43.774 | 42.218 | 9.728 |
| NAYARIT | 16.188 | 16.449 | 4.178 | 4.700 | 0.261 | 23.760 | 3.394 | 4.439 | 20.888 | 2.350 | 2.350 | 30.548 | 41.253 | 45.431 | 9.922 |
| NUEVO LEÓN | 10.250 | 11.597 | 1.594 | 1.374 | 0.879 | 14.702 | 2.198 | 3.023 | 7.227 | 0.989 | 0.632 | 11.761 | 49.217 | 40.918 | 5.001 |
| OAXACA | 15.768 | 12.656 | 3.631 | 5.394 | 1.141 | 16.183 | 3.423 | 3.008 | 25.519 | 1.971 | 2.697 | 33.299 | 43.050 | 44.295 | 9.232 |
| PUEBLA | 14.850 | 16.630 | 2.948 | 3.281 | 1.001 | 16.407 | 2.169 | 3.504 | 23.637 | 2.503 | 3.003 | 31.535 | 40.712 | 48.498 | 8.343 |
| QUERÉTARO | 11.736 | 14.425 | 3.423 | 2.689 | 0.978 | 17.359 | 4.890 | 3.178 | 17.604 | 1.711 | 2.200 | 28.362 | 47.677 | 38.631 | 6.846 |
| QUINTANA ROO | 11.429 | 15.238 | 1.270 | 0.635 | 1.905 | 9.206 | 2.857 | 2.222 | 23.492 | 6.032 | 4.762 | 38.413 | 51.111 | 30.476 | 4.127 |
| SAN LUIS POTOSÍ | 12.211 | 11.469 | 2.723 | 4.785 | 1.650 | 16.007 | 2.805 | 3.548 | 16.089 | 2.063 | 1.815 | 24.340 | 48.102 | 35.066 | 9.901 |
| SINALOA | 12.863 | 19.911 | 2.218 | 3.549 | 0.641 | 21.883 | 2.661 | 3.401 | 19.961 | 2.366 | 1.676 | 32.134 | 40.069 | 48.053 | 9.463 |
| SONORA | 13.734 | 18.842 | 2.043 | 2.838 | 2.270 | 23.156 | 2.724 | 5.675 | 20.318 | 1.476 | 1.022 | 29.512 | 48.354 | 41.317 | 7.151 |
| TABASCO | 15.422 | 21.687 | 1.205 | 2.088 | 1.526 | 20.321 | 1.124 | 2.972 | 13.976 | 3.614 | 1.687 | 21.365 | 42.570 | 50.361 | 5.221 |
| TAMAULIPAS | 13.875 | 15.107 | 2.381 | 1.888 | 1.888 | 18.062 | 2.545 | 3.530 | 9.278 | 1.642 | 0.903 | 16.256 | 49.015 | 42.529 | 5.008 |
| TLAXCALA | 14.211 | 10.702 | 2.456 | 2.807 | 0.877 | 15.088 | 3.684 | 2.632 | 18.947 | 4.211 | 1.930 | 22.982 | 43.333 | 45.789 | 7.544 |
| VERACRUZ | 13.582 | 15.734 | 2.965 | 3.252 | 1.387 | 18.747 | 3.396 | 2.965 | 18.890 | 2.726 | 1.435 | 31.707 | 46.389 | 42.037 | 8.082 |
| YUCATÁN | 10.733 | 16.718 | 1.651 | 2.786 | 1.342 | 18.885 | 1.238 | 2.683 | 13.829 | 3.096 | 2.167 | 22.910 | 47.678 | 40.454 | 7.637 |
| ZACATECAS | 14.390 | 16.211 | 4.007 | 5.100 | 1.821 | 25.137 | 4.736 | 4.918 | 23.133 | 2.186 | 1.457 | 27.687 | 41.712 | 41.894 | 14.026 |

\* Dirección General de Epidemiología, Secretaría de Salud (General Direction of Epidemiology, Health Ministry)
\*\* Instituto Nacional de Salud Pública, Encuesta Nacional de la Dinámica Demográfica (National Public Health Institute, National Survey of Demographic Dynamic)
\*\*\* Estimations obtained by CONEVAL (Consejo Nacional de Evaluación de la Política de Desarrollo Social, National Council for the Evaluation of Social Development Policy) based on the ENIGH (National Survey of Household Income and Expenditure) 2008, 2010, 2012, 2014, 2016, and 2018.
+ Instituto Nacional de Estadística y Geografía (National Institute of Statistics and Geography)
++ Consejo Nacional del Agua (CONAGUA) (National Water Commission)
+++ Instituto Nacional de Salud Pública (INSP) (The National Institute of Public Health of Mexico)

Additional variables concerning socio-demographic, economic, mobility, and climatic features were obtained at a state-level. In terms of socio-demographic variables, from the National Institute of Geography and Statistics (INEGI), we extracted information concerning clustering of individuals: population density (people per *km*^2^) in 2015 and the proportion of people in a household living in an overcrowded place in 2017; literacy rate of population aged ≥15 years in 2015; people settled in rural areas in 2010 (%) (a location was considered as rural when there are <2,500 habitants); and the number of physicians available by every 1000 people in 2015, which was obtained from the National institute of Public Health (INSP). In terms of economic variables, we obtained from INEGI the state contribution to gross domestic product (GDP) in 2018, which we modified by considering only those values associated with states not containing the biggest cities in Mexico to improve the linearity assumption with the transformed response. We also obtained information concerning people living in poverty in 2018 (%), as it is defined and calculated by CONEVAL according to a multidimensional index obtained from *per capita* income and an index of social deprivation (15). In terms of mobility, we extracted from INEGI information concerning internal migration, as the rate of people aged 5 years and over living in another state five years before 2014, and external migration, as the rate of people aged ≥5 years living in another country five years before 2014, both proxies of internal and external mobility, respectively; and, the number of flights in 2019 by state, which we calculated from information associated with the number of flights by airport in Mexico (Ministry of Communication and Transport)(16). Finally, information concerning average temperature (°C) in March 2020 was obtained from the National Council of Water (CONAGUA).

The risk of dying in tested individuals (t-CFRs) was chosen as the dependent variable on the basis of relevant indicators of COVID-19 epidemiology in Mexico; notably, the number of tests per 100,00 individuals is limited compared to other countries, which decreases detection rates and given the likely under-detection of mild SARS-CoV-2 cases in this setting, standardizing deaths by tested cases considers the extent of detection, which could similarly be influenced by structural factors (17). The remaining variables obtained and calculated from the different data sources are treated as explanatory, except for hospitalization, ICU, and intubation, which we considered as control variables, being an approximate measure of the presence of severe COVID-19 cases and possibly access to services attending COVID-19 in a region.

### COVID-19 t-CFRs estimation by state

We obtained quantile maps associated with raw and smoothed t-CFRs of COVID-19 cases. The risks by state were smoothed by using an empirical Bayes estimator, which is a biased estimator that improves variance instability proper of risks estimated in small-sized spatial units (18); however, the analyses were performed with both raw and smoothed risks to compare results. We also obtained maps concerning relative risks, understanding them as a comparison of the observed number of events by state to a national standard, the latter using the expected number of events considering as if risks in a state were the same as those at a national level.

### Spatial weight estimation and spatial autocorrelation

We obtained queen contiguity weights (19), which consider as neighbors those states sharing at least a point in common, obtaining a squared matrix of dimension 32 with all entries equal to zero or one, where a one indicates that two states are neighbors. From these neighbors, weights are calculated by integrating a matrix in a row-standardized form. Moran’s I statistic (20) was obtained as a measure of global spatial autocorrelation and its significance was assessed through a random permutation inference technique based on simulations. Local indicators of spatial autocorrelation (LISA) were obtained (21) and used to derive significant spatial clustering through four cluster types: High-High, Low-Low, High-Low, and Low-High. For instance, the Low-Low cluster indicates states with low values of a variable that are significantly surrounded by regions with similarly low values.

### Spatial multivariable linear model

A multivariable Generalized Geographically Weighted Regression (GGWR) was fitted (22). In this model, a dependent variable is measured for each spatial unit and independent variables (inputs) are simultaneously considered as well, such that the corresponding parameters depend on the coordinates in which the state is spatially located (centroids in the case of polygons); therefore, a parameter is associated with each state and independent variable. To be able to estimate such model, a weighting diagonal matrix is considered, we used Gaussian spatial weighting to generate it. These weights determine the relationship from any state to another in terms of the distance (Euclidean) between states and a bandwidth. The bandwidth determines which spatial units are similar under the GGWR and can be selected using automatized methods, we used a cross-validation (CV) method with an adaptive scheme, i.e. a different bandwidth was used for each unit. Since the response variable is a count (number of deaths), a GGWR with a Poisson distribution and logarithm link function was used, including as offset term the number of people tested with COVID-19 in a logarithmic scale, thus modelling the t-CFRs instead of just the number of deaths.

A global multivariable model for all states, a Poisson multivariable linear model (generalized linear model or GLM) with offset and a logarithmic link function, was also fitted and significant variables were identified (23). To obtain the best possible model, including variables with the greatest effect on the risks and satisfying as much as possible all statistical assumptions, we used the following selection process: 1) We fitted univariable Poisson models with offset and logarithmic link function, identified significant effects, and ordered them in absolute value from highest to lowest, 2) We identified variables with good and acceptable linear association with the log-transformed mortality risk by obtaining scatter plots between variables and the transformed risk, including a smoothed LOESS (locally estimated scatterplot smoothing) curve; and, when possible, variables were transformed to improve this assumption, as for GDP as explained above, 3) VIF was used to asses multicollinearity; thus, we fitted a model including variables with acceptable and good linear association, and eliminated any variable with a VIF>10, 4) We added to the resulting model one by one significant variables in the univariable models. First, we added the variable with the highest effect and fitted the associated model identifying whether VIF>10. If not, we added the variable, and if VIF>10, the model was not modified. We proceeded with the resulting model repeating the same process with the second highest effect; and so on, 5) From this process, we ended with a model consisting on all variables with the greatest univariable effects (0.08 and above in absolute value), better linear behavior, and without multicollinearity (VIF<=10), 6) We used residuals associated with other three Poisson models concerning ICU, intubated, and hospitalization, including a subset of variables pertaining to the model for fatality risk as inputs, thus obtaining the effects each confounder has over the risks without the effects that explanatory variables have over these confounders. These residuals and the corresponding estimated parameters do not have a meaning, 7) We evaluated goodness-of-fit and validated all model assumptions. For age, we obtained age groups: 0-1, 2-9, 10-19, …, 80-89, and 90 years old and over, fitted the corresponding univariable Poisson models with offset, and identified significant age groups and the direction of the association to obtain a new set of age groups: 10-39, 40-69, and 70 and more; and proceeded with these variables as with the others.

The GGWR with the same variables as in the GLM was fitted and multiplicative effects over the t-CFRs (i.e. the exponentiated estimated parameters) associated with each variable were calculated. Maps associated with these effects were obtained, presenting only those associated with the explanatory variables that were significant in the global model. All statistical analyses were conducted using R version 3.6.2 through the *spdep, rgdal*, and *spgwr* packages for spatial analyses and *car* package for the correlation analysis and GeoDa 1.14.0 was used also for some spatial analyses. The significance level for all analyses was 5% (i.e., alpha=0.05).

## RESULTS

### t-CFR description and spatial autocorrelation of COVID-19 t-CFRs between states

Maps for quartiles corresponding to the raw and smoothed t-CFRs are similar, except for two states, Tlaxcala, which from a category using the raw risks moves to the next superior one in the smoothed risks, the opposite occurring for Veracruz. Thus, only the map concerning the smoothed values is shown (**Figure 1A**). We observed the largest t-CFRs (above 4%) in Quintana Roo, Baja California, Chihuahua, and Tabasco, and the greatest relative risks (2.00-4.00) in Baja California, Chihuahua, Quintana Roo, and Tabasco (**Figure 1B**). Globally, there is a non-significant spatial autocorrelation (Moran’s I=-0.079, p=0.390); however, there is a noticeable Low-Low cluster on the northeast around San Luis Potosi and two Low-High clusters around Yucatan and Sonora (**Figure 2**). These Low-High clusters make sense since the associated states, especially Sonora, are surrounded by the states with the highest t-CFRs in all the country. We verified that the spatial autocorrelation value and clustering were exactly the same using both the raw values or those obtained with the Bayes spatial technique.

**Figure 1.**
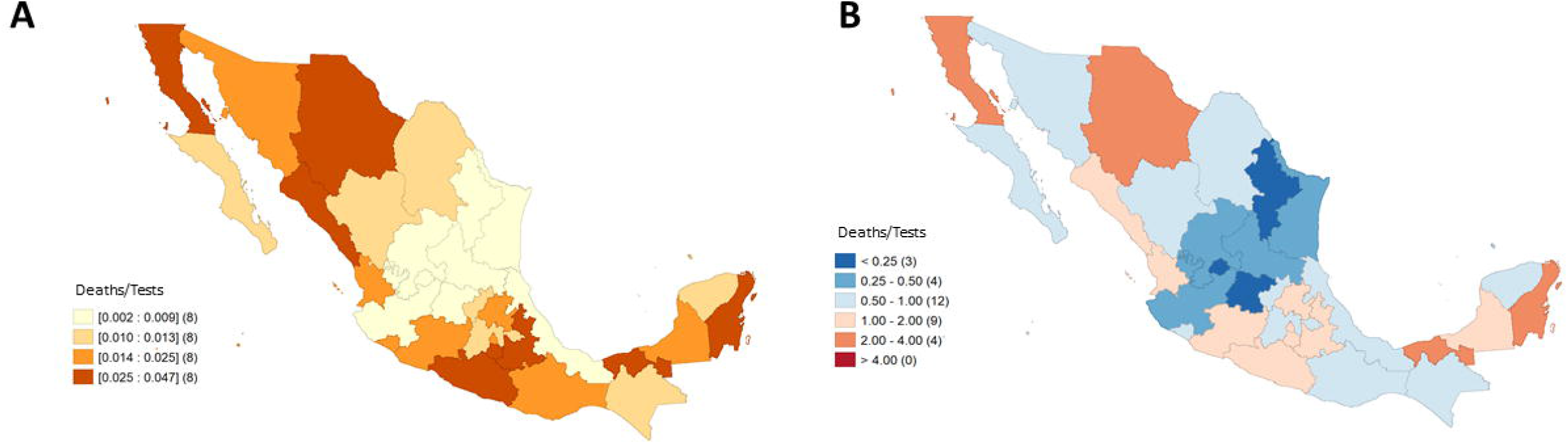
Maps associated with COVID-19 deaths in Mexico by state until April 21st, 2020. A) Quartiles corresponding to tested case-fatality risks smoothed through an empirical Bayes procedure. B) Relative risk.

**Figure 2.**
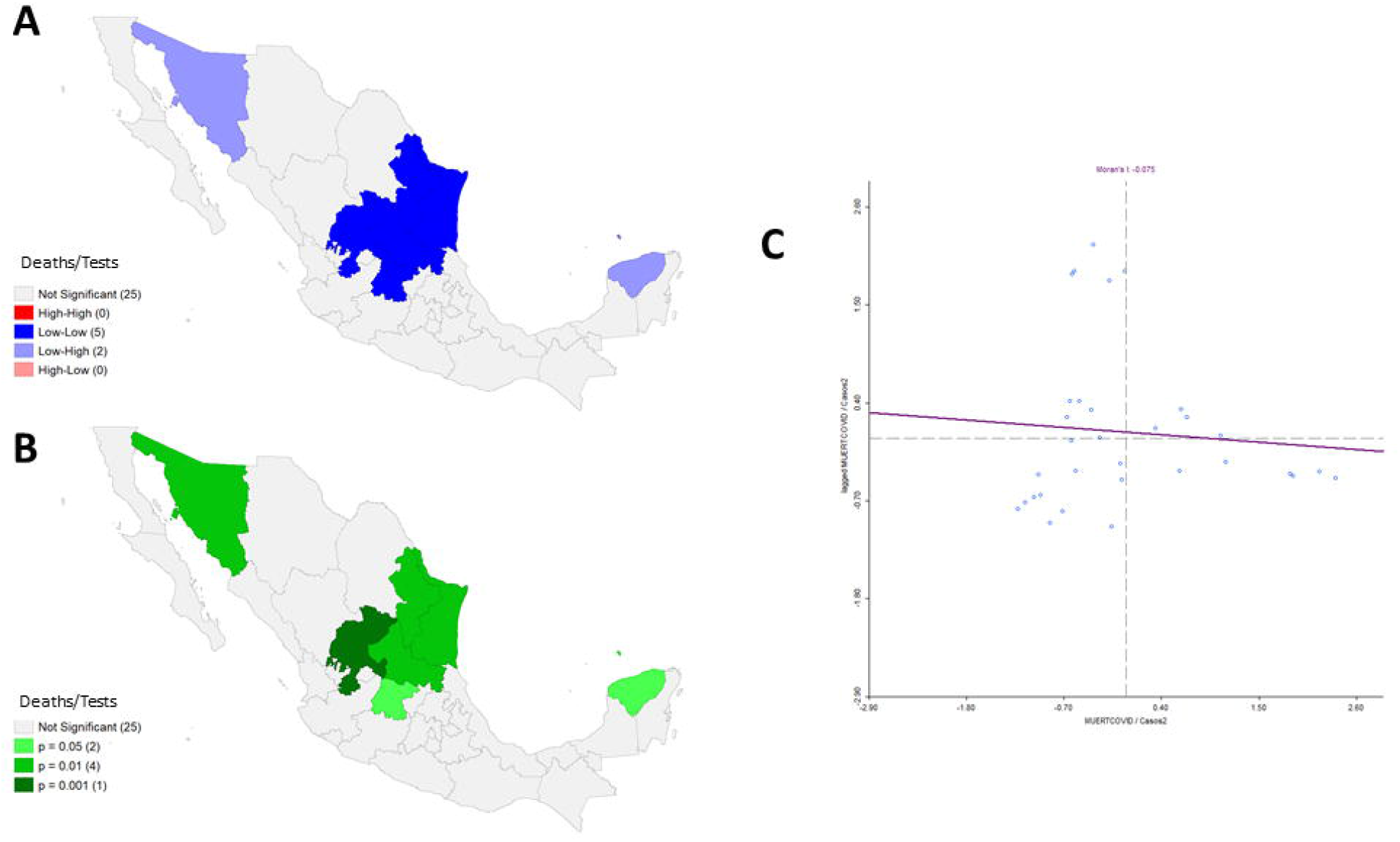
Spatial clustering associated with tested case-fatality risks of COVID-19 in Mexico by state until April 21st, 2020, considering queen contiguity. A) Significant spatial clustering obtained through Local Indicators of Spatial Autocorrelation (LISA) comparisons. There are four types of clusters: High-High, Low-Low, High-Low, and Low-High, e.g. a Low-Low cluster (blue) indicates states with low values of a variable significantly surrounded by regions with similarly low values. B) P-values associated with the spatial clustering in A), C) Scatter plot associated with the smoothed risks vs. their corresponding spatially lagged values, including the associated linear regression fitting, whose slope is the Moran’s I statistic.

### Fit of multivariable generalized global and geographical linear models for COVID-19 tested case-fatality risks

Through a preliminary analysis obtained by fitting a Poisson multivariable linear model with offset and including all variables, we found the presence of serious multicollinearity problems, since we obtained Variance Inflation Factors (VIF) with values above 50 for some variables. A correlation analysis between all variables was performed (**Figure 3A**), including the response (raw t-CFRs) identifying very correlated variables. Thus, we followed the model selection process as explained above. Our final global model included as inputs: diabetes, obesity, GDP, internal and external migration, age group of 10 to 39 years, physicians-to-population ratio, cardiovascular disease, ICU, hospitalization, and intubated. The latter three variables are confounders and to eliminate effects of other variables on them, they are used as residuals associated with appropriate Poisson models with independent variables: diabetes, obesity, age, physicians-to-population ratio, cardiovascular disease, plus ICU for the model associated with the intubated variable. Goodness-of-fit in our final model was assessed, finding that all variables were jointly significant (LR=472.19, p-value<0.001). Additionally, through a PP-plot associated with the standardized residuals and associated Anderson-Darling test (A=0.260, p-value = 0.689) and scatter plots between the fitted values and standardized residuals, and a similar plot using the root of the residuals instead, we determined that the link function and the way the explanatory variables are related with the response seems correct. However, we found some overdispersion (Chi-squared statistic divided by its degrees of freedom of 1.972). There were not any significant pairwise interaction effects.

**Figure 3.**
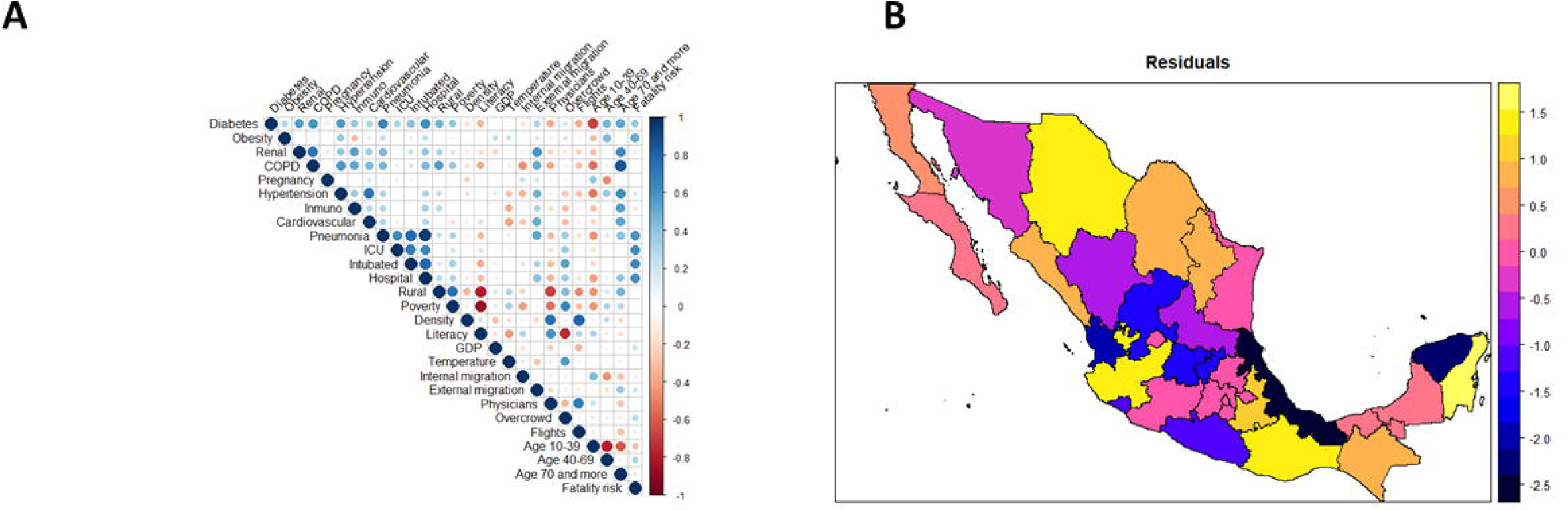
Figures associated with the selection and goodness-of-fit of a multivariable generalized geographically weighted model (GGWR); with a Poison distribution, offset, and a logarithm link function, used to explain tested case-fatality risks. A) Correlation plot including the raw risks, and B) Representation of Pearson residuals by state.

A multivariable GGWR with a Poisson distribution, adaptive kernel, and the same input variables was also fitted. In **Table 2**, we present a summary of the multiplicative effects over the risks, exponentiated parameters, i.e. minimum, quartiles, and maximum, associated with each variable for all states in the GGWR and the global values corresponding to the GLM. Pearson residuals associated with the GGWR are shown in **Figure 3B**, showing that the worst fit corresponded to two states: Veracruz and Yucatan.

**Table 2:** Statistics by variable (minimum, maximum, and quartiles) associated with the multiplicative effects by state over the tested case-fatality risks under the GGWR and analogous effects and p-values associated with a global model (all models consider a Poisson distribution, offset term, and logarithmic link function).

| Variable | Min | 1st quartile | Median | 3rd quartile | Max | Global |  |
| --- | --- | --- | --- | --- | --- | --- | --- |
|  |  |  |  |  |  | Effect | p-value |
| Intercept | 0.004 | 0.005 | 0.005 | 0.005 | 0.005 | 0.005 | <0.001 |
| Diabetes | 1.147 | 1.148 | 1.150 | 1.151 | 1.154 | 1.152 | <0.001 |
| Obesity | 1.121 | 1.122 | 1.123 | 1.125 | 1.125 | 1.122 | <0.001 |
| ICU (residual) | 1.219 | 1.220 | 1.220 | 1.222 | 1.225 | 1.223 | <0.001 |
| GDP (modified) | 1.298 | 1.298 | 1.298 | 1.300 | 1.307 | 1.304 | <0.001 |
| Internal migration | 1.250 | 1.252 | 1.262 | 1.268 | 1.276 | 1.267 | <0.001 |
| External migration | 1.322 | 1.323 | 1.325 | 1.332 | 1.361 | 1.349 | 0.018 |
| Age group 10-39 | 0.907 | 0.907 | 0.908 | 0.909 | 0.910 | 0.908 | <0.001 |
| Intubated (residual) | 1.214 | 1.216 | 1.219 | 1.221 | 1.222 | 1.219 | <0.001 |
| Physicians' ratio | 1.845 | 1.853 | 1.856 | 1.859 | 1.865 | 1.866 | <0.001 |
| Cardiovascular | 0.948 | 0.951 | 0.953 | 0.954 | 0.955 | 0.951 | 0.478 |
| Hospitalization (residual) | 0.957 | 0.957 | 0.957 | 0.959 | 0.960 | 0.958 | 0.009 |

### Predictors of COVID-19 spatial lethality in Mexico

The exponentiated estimated parameters under the GGWR for each state were obtained for each variable (not shown) and maps were obtained based on the .shp file provided in (24). Nevertheless, we only present the maps for those explanatory variables that significantly impacted over the log-transformed t-CFRs in the global model (GLM) (**Figure 4**). These significant variables were diabetes, obesity, GDP, internal and external migration, age group of 10 to 39 years, and physicians-to-population ratio. However, care should be taken when the map for GDP is interpreted considering this variable corresponds to GDP on states not having the biggest cities, as an interaction term between GDP and a binary variable, thus having a fixed value of zero in four states, which are represented as blank spaces in the map. All estimated terms were interpreted by considering fixed values for all variables except the one being interpreted.

**Figure 4.**
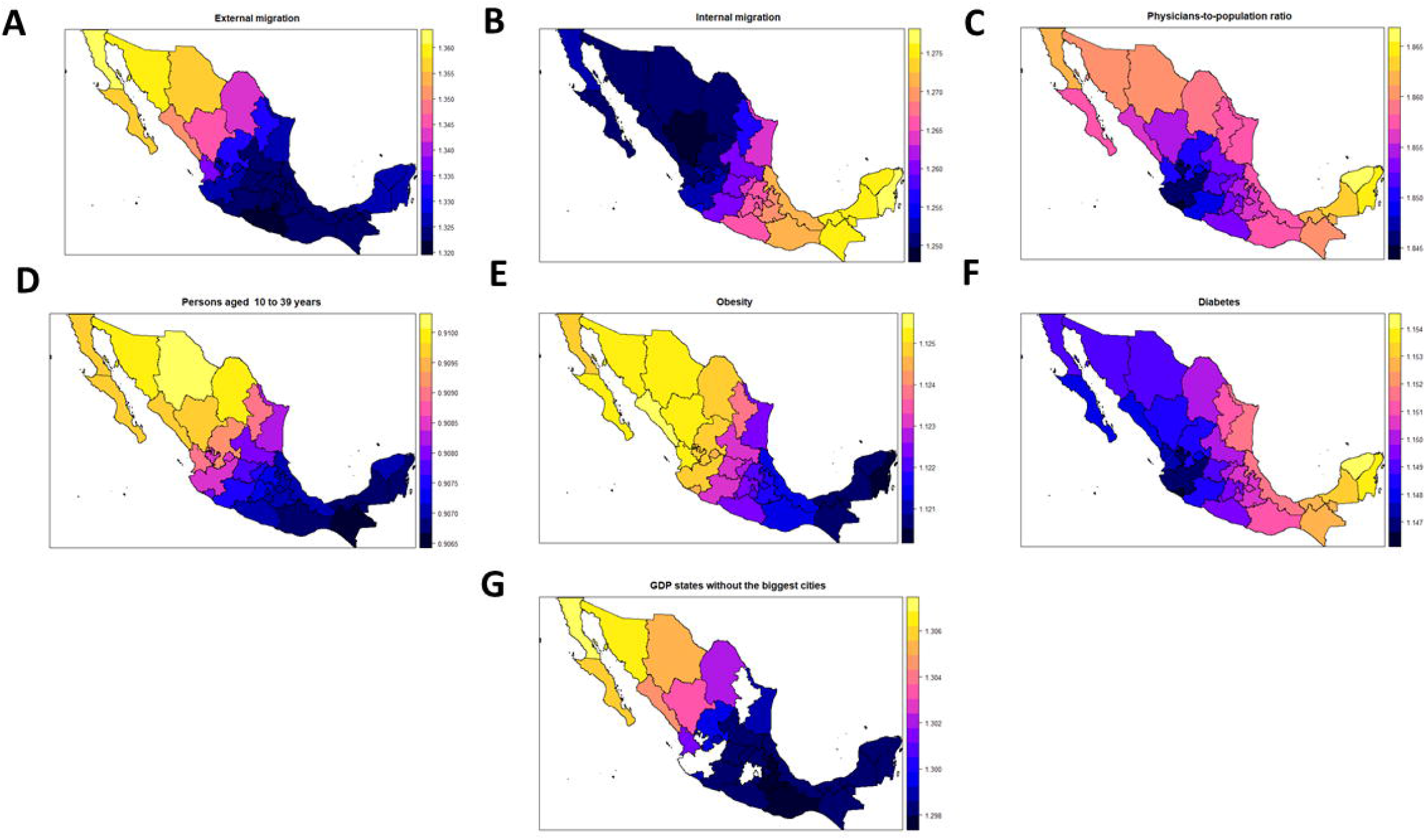
Multiplicative estimated effects over the tested case-fatality risks due to COVID-19 under a GGWR for those variables that significantly impact the response under a global model (Poisson models with offset and a logarithmic link function).

### Age and metabolic predictors of t-CFRs in Mexico

Prevalence of obesity has a global (for all states) significant positive association (multiplicative effect) with the COVID-19 t-CFRs (1.122; 95%CI 1.081-1.166), locally the effect is between 1.120 in Quintana Roo to 1.125 in Sinaloa, having a similar effect on all Mexico, though slightly larger on the north and center. For diabetes, there is also a significant positive association with the t-CFRs (1.152; 95%CI 1.079-1.230) with local effects between 1.147 in Colima to 1.154 in Yucatan, having a positive effect in all Mexico; but specially in the center, south, and Yucatan peninsula. On the other hand, the proportion of individuals between 10 and 39 years old has a significant negative association with the COVID-19 t-CFRs (0.907; 95%CI 0.873-0.944), locally the effect is between 0.907 in Chiapas to 0.910, thus having a similar association in all the country.

### Mobility and socio-economic predictors of t-CFRs in Mexico

The percentage of internal migration in the spatial unit a patient comes from has a significant positive association with the COVID-19 t-CFRs (1.267; 95%CI 1.183-1.356), locally the effect is between 1.250 in Durango to 1.276 in Quintana Roo having more effect in the south, center, and Yucatan peninsula. The percentage of external migration in the spatial unit a patient comes from is also significantly positively associated with the COVID-19 t-CFRs (1.349; 95%CI 1.052-1.728), locally the effect is between 1.322 in Morelos to 1.361 in Baja California, this effect exists over all Mexico, but it is stronger in the North and Baja California peninsula. Physicians-to-population ratio is also significantly positively associated with the t-CFRs (1.866; 95%CI 1.625-2.142), with local effects between 1.845 in Colima to 1.865 in Yucatan, having a slightly larger effect on the north and Yucatan peninsula. Finally, GDP excluding the states having the biggest cities in the country (Nuevo León, México, Ciudad de México, and Jalisco) is also significantly positively associated with the t-CFRs (1.304; 95%CI 1.198-1.429), locally the effects are between 1.298 in Oaxaca and 1.307 in Baja California.

## DISCUSSION

Here, we show the relevance of spatial analyses to allow us not only to understand how t-CFRs are distributed along Mexico and the presence of spatial clusters related to COVID-19 t-CFRs, but also how some variables are associated with these risks, with an association that variates along all territory. Through our analysis, we were able to identify spatial units or regions in which care could have been considered to avoid SARS-CoV-2 spread and its related adverse outcomes. Identifying these areas, might allow to understand the propagation of the disease in other possible waves of SARS-CoV-2 spread or similar infectious pathogens.

Considering the global results, variables that are significantly associated with an increase on the COVID-19 t-CFRs include percentages associated with diabetes, obesity, external and internal migration, physicians-to-population ratio, and GDP in states that do not include one of the greatest four cities in Mexico, whereas the proportion of individuals between 10 to 39 years old is significantly associated with a decrease on the risks. The association of cardio-metabolic diseases with adverse COVID-19 outcomes has been well documented and has been linked to mild, but sustained chronic inflammation, which may synergize with the cytokine storm associated with severe SARS-CoV-2 infection (25). Regarding other type of factors, internal and external migration have a very strong association with an increase of COVID-19 mortality, being external migration the variable with the second highest impact. This phenomenon is very particular for infectious diseases where greater movements of people move a disease from a geographical zone to another, which has occurred for instance for *Mycobaterium tuberculosis* or HIV/AIDS (26–28). The other two variables are related to urbanization, density, and economic importance of the states, specially physicians-to-population ratio. From the correlation plot, it can be seen that there is a very large positive association between physicians-to-population ratio and population density and overcrowded households, and a large negative association between this variable and both the rural and poverty proportions by state. On the other hand, the positive association between GDP and the fatality risks might also be related with states in which, due to the importance of economic activity, mobility could not be stopped after self-isolation measures, but that also do not have the health infrastructure that those states with the biggest cities have, in which, in spite of having many cases, there were less deaths.

The greatest t-CFRs, both raw and smoothed, corresponded to the states of Chihuahua, Quintana Roo, Baja California, and Tabasco (raw values of 5.11%, 4.79%, 4.34%, and 4.25%, respectively). It is noticeable how the relative risks in all these states have values between 2.5 and 3, suggesting that in these states the risks are above of what is nationally expected. We observed the presence of a spatial cluster concerning states with low risks; however, at least till the date associated with our dataset, there was no important clustering of states with high risks using the LISA technique.

The fact that Quintana Roo is an important touristic center explains the presence of COVID-19 cases. However, according to our models, the elevated t-CFR is mostly associated with the presence of diabetes and internal migration, besides of the physicians-to-population ratio, whose possible interpretation was discussed above. In Chihuahua and Baja California, the variables with a particular positive association with t-CFR were external migration, obesity, and GDP, whereas in Tabasco these variables correspond to diabetes and internal migration.

Globally, physicians-to-population ratio, which is heavily associated with more urbanity, overcrowding households and population density, and with less poverty, has the highest positive effect on the t-CFRs, though relatively similar on all Mexico. External migration has the second highest association with the t-CFRs particularly in those states in the north in which the risks were the highest, whereas internal migration has the fourth highest association, after GDP, with particular importance in the center, south, and Yucatan peninsula. The effect of obesity and diabetes over the risks is similar; for obesity, there is relatively a similar impact on all the country, whereas for diabetes there is a little higher effect on the center, south, and Yucatan peninsula. Age between 10 and 39 years old was the only variable associated with a less risks, which agrees with previous results analyzed using Mexican data, having a relatively similar effect on all the country (4,10).

Economic effects, such as poverty, may be better studied in a disaggregated model including it as a measure at an individual level. Unfortunately, such information is unavailable in the epidemiological data set, and at most, information concerning poverty at a municipality level could be attached to each individual, being state the spatial unit containing municipalities. However; we would still be using aggregated values and in the methodology used to calculate poverty in Mexico, the state values are estimators obtained from a representative sample, whereas the municipality values are estimated through small area estimation techniques; thus, being the former more reliable. In this sense, all analyses were performed at a state instead of at a municipal level. The reason behind this decision is that there were a lot of municipalities with zero values in the early time point we chose to focus our analysis, in both the number of COVID-19 cases and mortality and modelling such information with the probability distributions available for GGWR and other spatial linear models would not be possible. To obtain similar results we would require different tools, as zero-inflated geographical models. This could be possible future research work, including the identification of what are the characteristics of those municipalities with zero cases of the disease or no mortality. Additional future work corresponds also to analyze the data from a spatio-temporal framework, thus, the data set could be updated and the risks and associated factors followed through time for each state.

Our results are robust in terms of the model since it fits the data well and most of the statistical assumptions were satisfied; and, though there is some overdispersion, after fitting a quasi-Poisson linear model, we obtained the same results except that external migration was now significant at a 0.1 level. In terms of confounders, we fitted models with these variables as they are, as residuals, and as the first component in a Principal Component Analysis, finding in all cases that obesity, GDP, internal migration, age group of 10 to 39 years, and physicians-to-population ratio were significant.

In terms of the response, we obtained risks with the projected population in 2020 as denominator (population-fatality or mortality risks), and obtained similar results (maps) as the ones we present here, in terms of which states are on the highest quantiles, and in terms of similar clustering (29). Of notice is that using the projected values, we found a High-High cluster we did not find before in the Baja California peninsula. In fact, through the use of the Kulldorff’s spatial scan statistic (30), we also identified a cluster there, including three additional states (Sonora, Chihuahua, and Sinaloa); thus, it seems that a cluster in the north of higher risks exists, though it was not identified using the LISA methodology. Linear models using the projected values equivalent to the ones we fitted provided also similar results. For instance, in a model including the same variables as in our final model, we found the same variables significantly associated with the risks and direction of the association, except for diabetes, which was no longer significant. Analyses at an individual level were also fitted, using generalized (logistic) linear mixed models with a random intercept for state, and including mostly health related variables since socio-economic features are only at a state-level. We found that hospitalized status should be used separately from ICU and intubated, and that after taking care of multicollinearity, the results were similar as the ones at a state-level, in terms that variables significantly associated with the risks were still obesity, diabetes, and age group of 10 to 39 years, plus hypertension and cardiovascular.

It is important to notice that we are studying fatality risks associated with those individuals tested for the disease (t-CFRs); thus, care should be taken if results want to be extrapolated. As mentioned above, by using the projected population in 2020, instead of the tested individuals to model the mortality (population-fatality) risks, we obtained similar significant associations between the variables and risks, except for diabetes. However, we think this analysis is somewhat inaccurate in the sense that all health-related variables correspond to prevalence in individuals in the data set, which do not necessarily agree with those in the population, the same for age, and if we used population values by state, we would waste all the information in the epidemiological data, except for the number of deaths. Another option could have been to study case-fatality risks, thus considering the number of deaths and people infected; however, a similar problem arises, the real number of people with the disease is much larger than those being tested and results cannot be extrapolated, and this type of analysis is out of the scope of this study. Additionally, in any analysis, the number of infected and/or deaths are even more poorly estimated when analyzing the early spread of the disease, particularly in Mexico, in which not enough population had been tested. Despite these limitations, we were able to identify some spatial predictors of fatality risks associated with COVID-19 at an early stage of the pandemic, likely reflecting factors which could have been addressed to mitigate SARS-CoV-2 spread.

In conclusion, metabolic diseases, internal and external migration, physicians-to-population ratio, GDP per capita in states without the biggest cities, and age group between 10 to 39 years old were significantly associated with early COVID-19 fatality risks in Mexico. These predictors likely influence the growth of the pandemic moving forward, but variables as prevalence of metabolic diseases cannot be easily modified in the short-term. However, the identification of important variables in Mexico associated with the risks and in specific geographical areas, could help to decide necessary public policies which could have long-term impacts on future epidemic scenarios. Even though, this is an analysis in an early stage of SARS-CoV-2 spread, it allows us to understand how the pandemic evolved within Mexico and the possible measures that should be addressed for additional waves or similar diseases in Mexico and in specific zones of the country.

## Data Availability

Data included in the manuscript

## Author contributions

Conceptualization, methodology, formal analysis, validation, and supervision: RRA. Software and data curation: OYBC and RRA. Investigation and resources: JCGV and RRA. Analyses interpretation and writing (original draft, review, and editing): CGP, JCGV, OYBC, and RRA. Visualization: OYBC and RRA.

